# Safely managed sanitation practice and childhood stunting among under five years old children in Myanmar

**DOI:** 10.1101/2023.08.14.23294071

**Authors:** Than Kyaw Soe, Wongsa Laohasiriwong, Kittipong Sornlorm, Roshan Kumar Mahato

## Abstract

In 2020, 149 million children under the age of five were estimated to be stunted globally. Around half of deaths among children under 5 years of age are related to under-nutrition. Objective of this study is to determine the association between safely managed sanitation and childhood stunting among under-five years old children in Myanmar. This cross-sectional analytical study was conducted in 16 townships across three regions and five states in Myanmar. Multiple logistic regressions analysis was performed to determine the associations. This study found that 327 (27.09%) under-five children were stunted among a total of 1207 children in Myanmar. Children with unsafely managed sanitation were 2.88 times more likely to be stunting compared with children who access to safely managed sanitation services (AOR=2.88, 95% CI: 2.16 to 3.85; p-value <0.01). Other associated factors for childhood stunting were needs 1-15 minutes for water collection (AOR=2.07, 95% CI: 1.46 to 2.94; p-value <0.01), 15-60 minutes for water collection times (AOR=1.55, 95% CI: 1.08 to 2.23; p-value 0.02), unproper waste water disposal (AOR=1.99, 95% CI: 1.47 to 2.70; p-value <0.01), boys children (AOR=4.49, 95% CI: 3.30 to 6.12; p-value <0.01), did not take vitamin A supplements(AOR=1.64, 95% CI: 1.22 to 2.20; p-value <0.01), mothers height shorter than 153.4cm (AOR=1.94, 95% CI: 1.45 to 2.58; p-value <0.01), and the lower minimal diet diversity (AOR=1.53, 95% CI: 1.11 to 1.87; p-value 0.02). More access to safely managed sanitation facilities, technical sharing for proper waste water disposal, promoting household water supply system, health promotion for children’s diet eating pattern, and regular support for Vitamin A supplementation are critical to reduce childhood stunting among children under the age of five in Myanmar.

## 1. Introduction

In 2020, 149 million children under the age of five were stunted globally. Undernutrition is responsible for more than half of all fatalities in children under the age of five [1]. Moreover, half of all stunted children under the age of five lived in Asia and Africa [2]. Although global malnutrition has declined gradually since 2000, rapid progress is required to meet the 2030 SDG target.

Malnutrition is a major risk factor for childhood death because it attracts infectious diseases, increases the frequency and severity of illness, and impedes disease recovery[3]. If a child is malnourished, he or she has a low chance of achieving modest scholastic and intellectual success in adulthood, as well as low production, falling social level, and generational difficulties. Furthermore, malnutrition is linked to a higher risk of school dropout, lower socioeconomic status, and destitution in a long term[4, 5].

Chronic malnutrition caused frequent bouts of acute diarrhea in children. Furthermore, diarrhea remains a leading cause of death among children under the age of five, accounting for nearly 8% of all deaths among children under the age of five worldwide in 2017. Infectious diseases such as cholera, diarrhea, dysentery, hepatitis A, typhoid, and polio are spread due to poor sanitation. It is estimated that over 280,000 people die each year from diarrhea due by inadequate sanitation[6].

In Myanmar, diseases connected to water, sanitation, and hygiene (WASH) are common. In the long run, nutritional status of children under the age of five is poor when compared to other regional countries. Furthermore, 10% of children under the age of five had diarrhea, and 3% had acute respiratory infection symptoms in the two weeks preceding the data collection period in 2015-2016 Myanmar Demographic and Health Survey[7]. According to UNICEF, WHO, and the World Bank’s joint child malnutrition estimate (JME) for 2022, 24.1% of children under the age of five are stunted, 7.4% are wasting, and 0.8% are overweight in Myanmar [8].

Although every country strives to meet SDG target of safely managed sanitation by 2030, there is no dedicated survey to examine the level of safely managed sanitation status in Myanmar. Though various studies have been undertaken to verify the relationship between sanitation behavior and malnutrition among under five children, the limited studies have been conducted to investigate the relationship between access to safely managed sanitation and stunting among under five children in Myanmar. As a consequence, this research study will be useful for future studies of Myanmar and other countries relating with under five children nutrition. The evidence-based recommendations in this study were useful to policymakers in planning on sanitation and nutrition promotion for childhood malnutrition reduction programs.

## 2. Materials and methods

### 2.1 Study design and participants

This study was carried out in 16 townships across three regions and five states in Myanmar. To cover all parts of Myanmar, Kachin, Kayin, Mon, Rakhine, Shan states and Magway, Mandalay, Ayeyarwady regions were included in this study. A cross-sectional analytical study was carried out to determine the association between safely managed sanitation practice and stunting in children under the age of five. The children in the study ranged in age from 6 to 59 months at the time of data collection.

The inclusion and exclusion criteria were met by the eligible sample participants. Inclusion participants were i) children aged 6 months to less than 5 years old at the time of data collection, ii) children whose parents granted informed consent to participate in this study, and iii) children who had lived in this area for at least a year. Exclusion criteria included i) a child suffering from major health problems such as cancer, HIV, etc., ii) a child with congenital disorders, iii) a child with any physical or mental disability, and iv) a child living with a single mother. The multiple logistic regression formula was used to calculate the sample size (Hsieh, Bloch, & Larsen, 1998). According to calculation result of this formula, 1207 children of aged under five was sample size of this study.

Samples were chosen using multistage sampling. First, 16 townships were chosen from eight targeted Myanmar states and regions. Then, using simple random sampling, 1 ward was chosen from the urban context, and 2 villages for rural setting were chosen from these townships. And the household numbers, together with the names of the household leaders, were obtained from the township’s year-end health count data. Following that, 1207 sample houses with children under the age of five were chosen using a systematic random selection technique. Finally, the height of selected children, their mothers and fathers were measured using conventional instruments, and their mothers were interviewed using preformed questionnaires.

### 2.2 Measures

Children’s body height in centimeters (cm) were measured to the nearest 0.1 cm by using metering object that was recognized and used in township health department. Stunting (height for age) was classified as the children whose height for age Z score was below minus two standard deviation (-2SD) than their expected height for age.

According to the WHO/UNICEF Joint Monitoring Program (JMP) definition, safely managed sanitation was defined as the as practicing the improved sanitation facilities by not sharing with other families in which excreta were disposed safely in situ or removed and providing treatment outside. At least basic drinking water was classified as drinking water from an improved source, provided collection time is not more than 30 minutes for a round trip, including queuing. Basic hygiene level was defined as the availability of a handwashing facility on premises with soap and water.

Child using toilet, rinsing feces into toilet and buried practices were considered as proper child feces disposal, but rinsing feces into drain, throw into garbage, left in open, use as manual were considered as unproper child feces disposal, Disposing garbage via formal service provider, informal services provider, dispose in designated locations, burying and burning were described as proper solid waste disposal, whereas disposing within own plot, dispose elsewhere and don’t know were regarded as unproper solid waste disposal. Drain to the sewer line, designated pit, and soaking pit were considered proper waste water disposal, whereas drain to septic tank, open ground, water body, and elsewhere were considered unproper waste water disposal.

Health literacy was assessed in the areas of sanitation promotion, hygiene promotion, and nutritional promotion. Health literacy score intervals were classified into four levels for each percentage: less than 60 percent as “inadequate,” 60 to 69 percent as “problematic,” 70 to 79 percent as “adequate,” and more than 80 percent as “excellent.”

There were 7 parts in question structures: 1) socio demographic factors, 2) child factors, 3) parent factors, 4) WASH factors, 5) food safety and diet pattern, 6) health literacy and 7) disease factors. In socio demographic factors, place of residence, kitchen location, having animal feces in house, house floor type and family size were included. Age, gender, parity, gestational age, birth weight, birth interval, Vitamin A supplement, deworming, antenatal care visit and exclusive breast feeding were included in child factor questionnaires. In terms of parent factors, age, height, education, occupation, income, smoking, nutritional knowledge and attitude were added. Relating with WASH factors, services level of water supply, sanitation and hygiene, systematic hand washing of mother, father and child, water collection time, water shortage, solid waste disposal, child feces disposal and waste water disposal were included. Food safety score, main food, food diversity and food avoidance information were included in food safety and diet pattern questions. Sanitation promotion, hygiene promotion and nutrition health literacy were included for health literacy parts. And diarrhea, dysentery, worm infection and sickness times were questioned for disease factors.

### 2.3 Statistical Analysis

Microsoft MS Excel was used to record the raw data of 1207 respondents. The data were inverted using Stata version 13.1. For categorical data, the participants’ socio-demographic and baseline characteristics were characterized using frequency and percentage, and for continuous data, mean, median, minimum, maximum, and standard deviation were presented. Multiple logistic regression, adjusted OR with 95% confidence interval, and pvalue were used to determine the association between safely managed sanitation and stunting. All test data were two-sided, and statistical significance was defined as a p-value less than 0.05.

## 3. Results

### 3.1 Prevalence of stunting among under five children

Stunting prevalence by their age and gender among under five children in Myanmar was presented in table 1. In a total of 1207 under-five children, 327 (27.09%) were stunted. The mean age Z-value (HAZ) for height was -1.10 (minimum -4.38, maximum 3.34). Stunting prevalence by age group is highest in children aged 2 to 3 years (33.62%), followed by children aged 4 to 5 years (32.38%), 6 months to 1 year (28.38%), 1 to 2 years (26.46%), and 3 to 4 years (15.69%). In terms of the prevalence of stunting by gender, 14.64 percent of all girls and 38.13 percent of all boys were stunted.

**Table 1.**
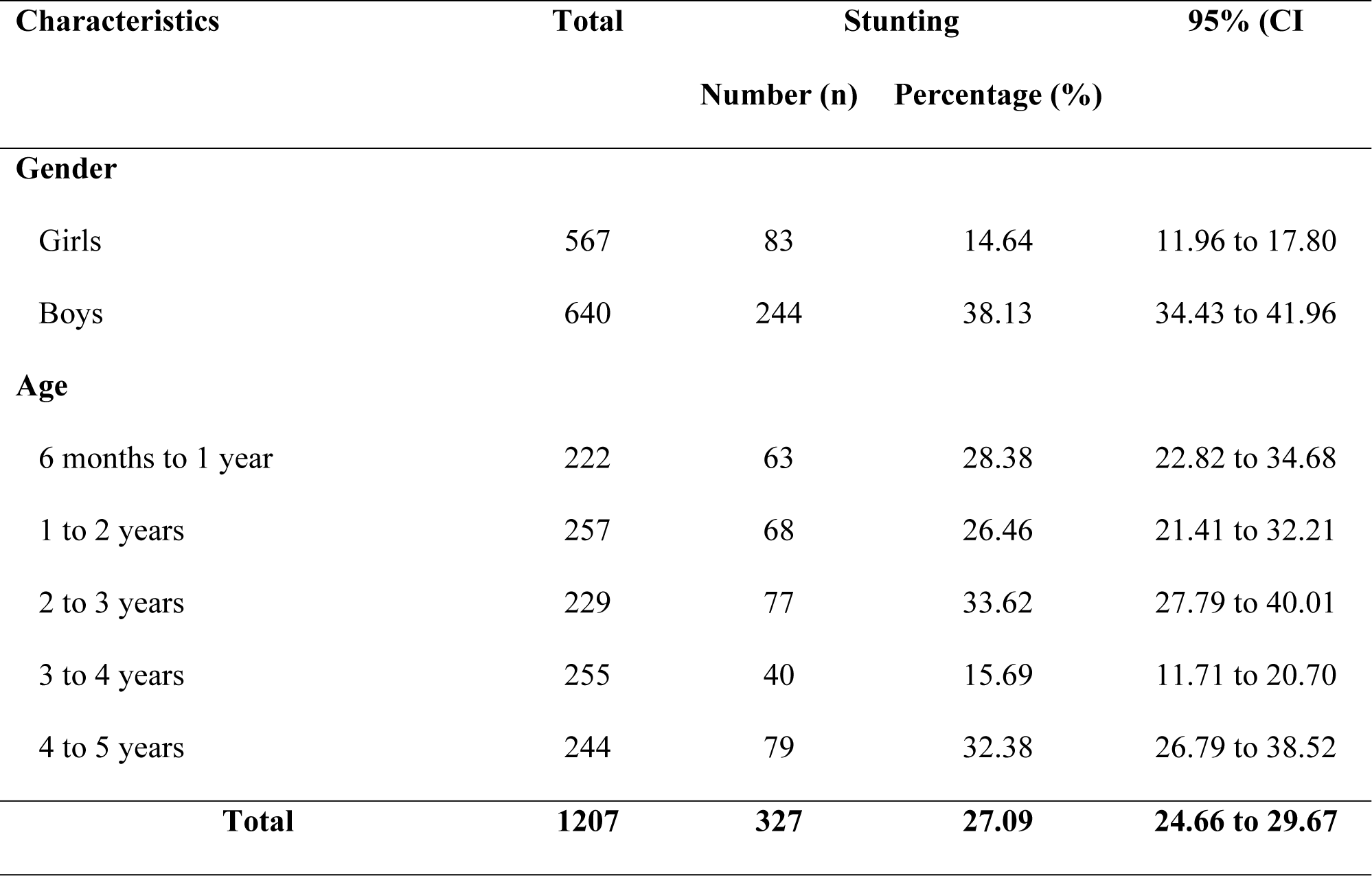
Prevalence of stunting by their age and gender among under-five children in Myanmar.

| Characteristics | Total | Stunting |  | 95% (CI |
| --- | --- | --- | --- | --- |
|  |  | Number (n) | Percentage (%) |  |
| Gender |  |  |  |  |
| Girls | 567 | 83 | 14.64 | 11.96 to 17.80 |
| Boys | 640 | 244 | 38.13 | 34.43 to 41.96 |
| Age |  |  |  |  |
| 6 months to 1 year | 222 | 63 | 28.38 | 22.82 to 34.68 |
| 1 to 2 years | 257 | 68 | 26.46 | 21.41 to 32.21 |
| 2 to 3 years | 229 | 77 | 33.62 | 27.79 to 40.01 |
| 3 to 4 years | 255 | 40 | 15.69 | 11.71 to 20.70 |
| 4 to 5 years | 244 | 79 | 32.38 | 26.79 to 38.52 |
| Total | 1207 | 327 | 27.09 | 24.66 to 29.67 |

### 3.2 Baseline characteristics of under five years old children

Table 2 showed the baseline characteristics of the respondents. Majority of them were from rural areas (78.62%). 84.59 percent of children’s kitchen located inside the home. The majority of respondents’ house floors were made of concrete or wood. Two-thirds of families had less than five members. In this survey, the boy-to-girl ratio was 47:53. Eighty percent of families had one or two children. 3.35 percent of babies were born prematurely, while 5.05 percent were born with low birth weight. Vitamin A supplement was taken by two-thirds of the children. 59.15 percent of all children received deworming medications, and 78.13 percent had antenatal treatment while their pregnant. The majority of child (76.22%) was treated with exclusive breast feeding.

**Table 2.** Baseline characteristics of the respondents.

| Characteristics | Total (n=1207) |  |
| --- | --- | --- |
|  | Number | Percentage |
| Place of residence |  |  |
| Urban | 258 | 21.38 |
| Rural | 949 | 78.62 |
| Kitchen location |  |  |
| Outside home | 186 | 15.41 |
| Inside home | 1021 | 84.59 |
| House floor type |  |  |
| Concrete or wood | 1023 | 84.76 |
| Earth or bamboo | 184 | 15.24 |
| Family size |  |  |
| < 5 members | 853 | 70.67 |
| ≥ 5 members | 354 | 29.33 |
| Mean (SD) | 4.08 (± 1.17) |  |
| Median (Min:Max) | 4 (3:11) |  |
| Gender of child |  |  |
| Boy | 567 | 46.98 |
| Girl | 640 | 53.02 |
| Parity |  |  |
| 1 – 2 children | 957 | 79.29 |
| 3 and more children | 250 | 20.71 |
| Labor type |  |  |
| Preterm labor (birth before 37 weeks) | 44 | 3.65% |
| Normal labor (birth after 37 weeks) | 1163 | 96.35 |
|  | Number | Percentage |
| **Birth weight (kg)** |  |  |
| Low birth weight (< 2.5 kg) | 61 | 5.05 |
| Normal birth weight (≥ 2.5 kg) | 1146 | 94.95 |
| **Birth interval (years)** |  |  |
| First child | 588 | 48.72 |
| Less than 3 years | 134 | 11.10 |
| Equal and more than 3 years | 485 | 40.18 |
| Mean (SD) | 30.69 (± 38.22) | |
| Median (Min:Max) | 20 (0 : 172) |  |
| **Vitamin A supplement** |  |  |
| No | 435 | 36.04 |
| Yes | 772 | 63.96 |
| **Deworming** |  |  |
| No | 493 | 40.85 |
| Yes | 714 | 59.15 |
| **Antenatal Care** |  |  |
| Yes | 943 | 78.13 |
| No | 264 | 21.87 |
| **Exclusive breast feeding** |  |  |
| No | 287 | 23.78 |
| Yes | 920 | 76.22 |
| **Mother's current age** |  |  |
| Under 30 | 526 | 43.58 |
| 30-40 years | 559 | 46.31 |
|  | Number | Percentage |
| Over 40 years | 122 | 10.11 |
| Mean (SD) | 30.86 | (± 6.03) |
| Median (Min:Max) | 30 | (15 : 51) |
| **Mother's height in cm** |  |  |
| Less than 153.4 cm | 594 | 49.21 |
| Equal and more than 153.4 cm | 613 | 50.79 |
| Mean (S.D.) | 154.19 | (± 5.11) |
| Median (Min: Max) | 154 | (145 : 195) |
| **Father's height in cm** |  |  |
| Less than 163.5 cm | 947 | 78.46 |
| Equal and more than 163.5 cm | 260 | 21.54 |
| Mean (S.D.) | 159.26 | (± 5.03) |
| Median (Min: Max) | 158 | (150 : 178) |
| **Mother's education** |  |  |
| Graduated | 122 | 10.11 |
| Non graduated | 1085 | 89.89 |
| **Mother's occupation** |  |  |
| Public or private employee | 122 | 10.11 |
| Other business or dependent | 1085 | 89.89 |
| **Father's occupation** |  |  |
| Public or private employee | 104 | 8.62 |
| Other business or dependent | 1103 | 91.38 |
| **Monthly family income in USD** |  |  |
| Less than 100 USD | 670 | 55.51 |
|  | Number | Percentage |
| Equal and more than 200 USD | 537 | 44.49 |
| Mean (SD) | 118.50 | (± 77.55) |
| Median (Min:Max) | 95 | (2 : 1286) |
| **Mother smoking** |  |  |
| No | 1182 | 97.93 |
| Yes | 25 | 2.07 |
| **Father smoking** |  |  |
| No | 687 | 56.92 |
| Yes | 520 | 43.08 |
| **Mother's knowledge on child malnutrition** |  |  |
| Good | 1143 | 94.70 |
| Poor | 64 | 5.30 |
| **Mother's attitude on child malnutrition** |  |  |
| Good | 932 | 77.22 |
| Poor | 275 | 22.78 |
| **Drinking water services level** |  |  |
| At least basic | 826 | 68.43 |
| Limited | 14 | 1.16 |
| Unimproved | 134 | 11.10 |
| Surface water | 233 | 19.30 |
| **Sanitation services level** |  |  |
| Safely managed | 719 | 59.57 |
| Basic | 80 | 6.63 |
| Limited | 221 | 18.31 |
|  | Number | Percentage |
| Unimproved | 100 | 8.29 |
| Open defecation | 87 | 7.21 |
| **Hygiene services level** |  |  |
| Basic | 1058 | 87.66 |
| Limited | 49 | 4.06 |
| No handwashing facility | 100 | 8.29 |
| **Water collection time** |  |  |
| No need to go for water collection | 677 | 56.09 |
| 0 to 15 minutes | 302 | 25.02 |
| 15 to 60 minutes | 228 | 18.89 |
| Median (Min: Max) | 18 | (2:29) |
| Median (Min: Max) | 0 | (0:60) |
| **Water shortage experience** |  |  |
| No | 1135 | 94.03 |
| Yes | 72 | 5.97 |
| **Child feces disposal** |  |  |
| Proper disposal | 1025 | 84.92 |
| Unproper disposal | 182 | 15.08 |
| **Solid waste disposal** |  |  |
| Proper disposal | 836 | 69.26 |
| Unproper disposal | 371 | 30.74 |
| **Waste water disposal** |  |  |
| Proper disposal | 532 | 44.08 |
| Unproper disposal | 675 | 55.92 |
|  | Number | Percentage |
| Overall food safety score |  |  |
| Less than 17 | 99 | 8.20 |
| 17 to 24 | 1108 | 91.80 |
| Mean (SD) | 19.67 (± 2.18) |  |
| Median (Min:Max) | 20 (12:24) |  |
| Main food |  |  |
| Rice and others | 1,110 | 91.96 |
| Breastmilk | 97 | 8.04 |
| Minimum dietary diversity at least 4 |  |  |
| Yes | 706 | 58.49 |
| No | 501 | 41.51 |
| Eating times per day |  |  |
| Less than 4 times | 850 | 70.42 |
| Equal and more than 4 times | 357 | 29.58 |
| Food avoiding |  |  |
| No | 974 | 80.70 |
| Yes | 233 | 19.30 |
| Sanitation promotion health literacy |  |  |
| Inadequate | 217 | 17.98 |
| Problematic | 263 | 21.79 |
| Adequate | 641 | 53.11 |
| Excellent | 86 | 7.13 |
| Hygiene promotion health literacy |  |  |
| Inadequate | 326 | 27.01 |
|  | Number | Percentage |
| Problematic | 283 | 23.45 |
| Adequate | 526 | 43.58 |
| Excellent | 72 | 5.97 |
| **Nutrition promotion health literacy** |  |  |
| Inadequate | 211 | 17.48 |
| Problematic | 298 | 24.69 |
| Adequate | 625 | 51.78 |
| Excellent | 73 | 6.05 |
| **Diarrhea in past two weeks** |  |  |
| No | 1121 | 92.87 |
| Yes | 86 | 7.13 |
| **Dysentery in past two weeks** |  |  |
| No | 1156 | 95.77 |
| Yes | 51 | 4.23 |
| **Worm infection in past two weeks** |  |  |
| No | 1159 | 96.02 |
| Yes | 48 | 3.98 |
| **Sickness in last month** |  |  |
| No | 651 | 53.94 |
| Yes | 556 | 46.06 |

43.58 percent of mothers were under 30 years of age, 46.31 percent were 30 to 40 years old and 10.11 percent were over 40 years old. Median height of mother were 154 cm and median height of father were 158 cm. One out of ten mothers were graduated. 10.11 percent of mothers and 8.62 percent of fathers worked as public or private employee. 55.51 percent earned less than 100 USD income per month. 2.07 percent of mothers and 43.08 percent of father were current smokers. 94.70 percent of moms had a good nutritional knowledge, and 77.22 percent of mothers had a positive attitude on nutrition.

In terms of water, sanitation and hygiene (WASH) characteristics, 68.43 percent were accessible for at least basic water supply services. Six out of ten families had safely managed sanitation services. Nine out of ten families were in basic hygiene level. 56.09 percent of all households do not require time to acquire water and can obtain it at home. During last year, 5.97 percent of people had a water deficit. 15.08 percent household improperly disposes of child feces. Three out of every ten houses improperly disposed of garbage, and 55.92 percent household improperly disposed of waste water.

The food security score ranged from 12 to 24, with 20 being the mean and median. Rice was the major diet for nine out of 10 children. 41.51 percent of children consumed at least four different types of foods. Some foods were avoided by 19.20 percent of all children. The majority of children’s mothers had appropriate health literacy scores, with 53.11 percent having adequate sanitation health literacy, 43.58 percent having adequate hygiene promotion health literacy, and 51.78 percent having adequate nutrition promotion health literacy. Before two weeks, 7.13 percent had diarrhea, 4.23 percent had dysentery, and 3.98 percent had worm infection. In the previous month, 46.06 percent children were reported unwell.

### 3.3 Factors associated with stunting among under five years old children

After bivariate analysis, house floor type, family size, gender of child, gestational age, vitamin A supplement, antenatal care, exclusive breast feeding, maternal height, paternal height, mother’s education status, family income, mother smoking, father smoking, maternal attitude on nutrition, water services level, water collection time, water shortage experience, child feces disposal, solid waste disposal, waste water disposal, food safety score, minimum diet diversity, sanitation promotion health literacy, nutrition promotion health literacy, diarrhea, dysentery, worm infection and sickness were selected to add on multivariate model as their analyzed pvalue was less than 0.25.

Table 3 present factors associated with stunting among under five years old children in Myanmar. After controlling the confounding factors with backward elimination multivariate analysis, safely managed sanitation was found as a strongly significant factor for childhood stunting among under-five children. In terms of other WASH factors, water collection times and waste water disposal were found as associated factors for stunting among under five years old children in Myanmar.

**Table 3.** Factors associated with stunting among under five years old children in Myanmar.

| Variable | Total<br>(No.) | Stunting (%) | Crude<br>OR | Adjusted<br>OR | 95% CI | P-value |
| --- | --- | --- | --- | --- | --- | --- |
| **Sanitation services level** |  |  |  |  |  |  |
| Safely managed sanitation | 719 | 19.89 | 1 | 1 |  |  |
| Non-safely managed sanitation | 488 | 37.70 | 2.44 | 2.88 | 2.16 to 3.85 | <0.01 |
| **Water collection time** |  |  |  |  |  |  |
| No need time | 677 | 21.12 | 1 | 1 |  |  |
| 1-15 minutes | 302 | 35.43 | 1.98 | 2.07 | 1.46 to 2.94 | <0.01 |
| 15-60 minutes | 228 | 33.77 | 1.83 | 1.55 | 1.08 to 2.23 | 0.02 |
| **Waste water disposal** |  |  |  |  |  |  |
| Proper disposal | 532 | 18.42 | 1 | 1 |  |  |
| Unproper disposal | 675 | 33.93 | 2.23 | 1.99 | 1.47 to 2.70 | <0.01 |
| **Gender of child** |  |  |  |  |  |  |
| Girl | 567 | 14.64 | 1 | 1 |  |  |
| Boy | 640 | 38.13 | 3.59 | 4.49 | 3.30 to 6.12 | <0.01 |
| **Vitamin A supplement** |  |  |  |  |  |  |
| Yes | 435 | 22.54 | 1 | 1 |  |  |
| No | 772 | 35.17 | 1.86 | 1.64 | 1.22 to 2.20 | <0.01 |
| **Mother's height in cm** |  |  |  |  |  |  |
| Equal and more than 153.4 cm | 613 | 22.19 | 1 | 1 |  |  |
| Less than 153.4 cm | 594 | 32.15 | 1.66 | 1.94 | 1.45 to 2.58 | <0.01 |
| **Diet diversity at least 4** |  |  |  |  |  |  |
| Yes | 706 | 22.95 | 1 | 1 |  |  |
| No | 501 | 30.03 | 1.44 | 0.53 | 1.11 to 1.87 | 0.02 |

Children who accessed to unsafely managed sanitation services meaning that practicing basic, limited, unimproved and open defecation were more likely to be stunting compared with children who access to safely managed sanitation services about 3 times (AOR=2.88, 95% CI: 2.16 to 3.85; p-value <0.01).

Moreover, those children whose family need to take 1 to 15 minutes for water collection (AOR=2.07, 95% CI: 1.46 to 2.94; p-value <0.01) were significantly more likely for 2 times to be stunted than the children whose family does not need to take time for water collection. Likewise, children whose family need to take more than 15 to 60 minutes for water collection had 55% more chances to be stunted than reference children (AOR=1.55, 95% CI: 1.08 to 2.23; p-value 0.02). Furthermore, children who live in a family with unproper waste water disposal practice were 2 times more likely to be stunted than the children with a family practicing proper waste water disposal methods.

Children’s gender, taking vitamin A supplement, mother’s height and minimum diet diversity were also the associated factors childhood stunting. Under five years old boys were 4.5 times more likely to be stunted than the girls aged under five (AOR=4.49, 95% CI: 3.30 to 6.12; p-value <0.01). Besides, children who did not take vitamin A supplement had 64% more chances to be stunting than other children who took vitamin A supplement (AOR=1.64, 95% CI: 1.22 to 2.20; p-value <0.01). Moreover, children who bore from mother with shorter than 153.4 cm were more likely to be stunted about 2 times than the children from mother with taller than 153.4 cm (AOR=1.94, 95% CI: 1.45 to 2.58; p-value <0.01). Furthermore, children who did not eat minimum diversity of diet at least 4 items were significantly to be stunted about 50% more than children who eat at least 4 diversities of foods.

## 4. Discussion

Childhood malnutrition is a critical health problem in Myanmar, which is one of the high malnutrition prevalence countries in South East Asia. This study found that 327 (27.09%) of 1207 children were stunted, which means that nearly three out of every ten children under the age of five were shorter than their median height for age of normal children. The prevalence of stunting of this study was slightly lower than the Myanmar Demographic Health Survey 2016 (MDHS 2016) result findings of 29 percent[7].

Safely managed sanitation practice was significantly associated with childhood stunting among under-five children in Myanmar after controlling for confounding factors with backward elimination multivariate analysis. Other two water, sanitation, and hygiene (WASH) variables such as water collection times, waste water disposal, and other four child-related factors such as children’s gender, taking vitamin A supplement, mother’s height, and minimum diet diversity were also associated with childhood stunting among children under the age of five in Myanmar.

Children who had access to safely managed sanitation services were three times less likely to be stunted than other children. The conclusions of this study supported the findings of the previous Indonesia study[9]. Indonesia study presented that ambient sanitation characteristics where children live, as well as ownership of a semi-permanent toilet, have a strong association with the occurrence of stunting. However, this new finding of association between safely managed sanitation and stunting supports the prior systematic review and meta-analysis findings of Rosiyati, Vilcin, and Larsen about the relationship between childhood stunting and sanitation status[10–12].

Momberg study on South Africa explored a relationship between safely managed sanitation and children’s nutritional health[13] of under one year old children. The current study findings support the previous conclusions on the association between safely managed sanitation and childhood stunting. Furthermore, this new finding on sanitation service level and childhood stunting contributed to the findings of India[14], Pakistan[15], Indonesia[16] and Ethiopia[17] on this association of sanitation and childhood stunting. Therefore, we need to focus for improving safely managed sanitation facilities among the community for decreasing children malnutrition.

Difficulty to get water and paying the time for water collection was also the risk factor for childhood stunting. This study identified that under five years old children from the home with no need to pay time for water collection can reduce the stunting from 8% to 3 times. Previous study[18] also found that two to five years old children from households with improved water source and pipe water system had lower opportunity to be stunting. Water can be contaminated through-out the water collection route such as water source, fetching, carrying, carrier material, etc.,. This finding supported to former study of Mashida who highlighted that children who use domestic water from surface water source such as river, stream, etc., were nine times more likely to be malnutrition than the children those from households with pipe water[19]. Therefore, these all findings highlighted the importance of household water accessible at home for child nutrition improvement.

This study explored that unproper waste water disposal was the predisposing factor for childhood stunting. Children from a family with proper waste water disposal practice had 2 times less likely to be stunting than the children with unproper waste water proposal practice. Waste water contained the dissolved matter and microorganisms that may be harmful to humans, animals and the environment. And unproper water way and poor drainage line were the source for fly and mosquito breeding which can cause the source of various infections. Frequent sickness and infection affect to the childhood nutritional status. There were no former studies on relationship between waste water disposal type and childhood nutritional status.

Gender of children was the associated factors for childhood stunting proved by many studies. This study found that boys were more likely to be stunting about 4.5 times than girls in Myanmar. This study result was same with findings of Tanzania[18], Nigeria[20] and Indonesia[21] studies, those studies found that male children were more prominent to be stunting than female. However, the odd of being stunted was about three times higher for Ethiopian females[22]. Indian girls had higher heigh for age Z scores than boys[23]. This finding fulfilled previous Myanmar literature of showing the association between gender and childhood stunting. That study revealed that boy had a higher risk to be stunted than girl in Myanmar[24].

This study found that children who took Vitamin A supplement were less likely to be stunted than other children. Uganda study also evidenced that Vitamin A deficiency was the predisposing factor for childhood stunting[25]. Similarly, one Iran national study identified the association between serum retinol level and toddler’s stunting[26]. This finding contributed to the former findings on relationship between Vitamin A supplement and children stunting. Regular Vitamin A supplement to under five children was taking an important role for decreasing stunting in children.

Previous Myanmar studies of Kang and Phyo proved that children from short mother were 2.5 to nearly 4 times more chances to be stunted than the children from tall mothers[24, 27]. Moreover, Li systematic review paper finding supported for this finding that short maternal height has 4 time to have stunted child than tall mother[28]. This study fulfilled the previous findings for this relationship. This study explored that children whose mother was shorter than Asian’s average woman height were more likely to be stunted about 2 times than the children from taller mothers.

This study also found that eating diversity of food was the significant factor determining for under five children stunting. Children eating minimum diet at least 4 items were less likely about 53% than children who eat diverse foods less than 4 items per day. Similar result was evidenced in one India study, presenting that lower consumption of varieties of foods such as grains, roots and tubers was associated to have 34% higher risk of childhood stunting[14]. Thes studies encouraged all 6 to 59 months old children to eat at least four varieties of food per day for their nutritional improvement.

### Strengths of Study

This study explored the prevalence stunting among children aged under five years old in Myanmar. It is the first report on finding the association between safely managed sanitation practice and under five children’s stunting in Myanmar. Therefore, this research study can be a reference for similar future studies which will be conducted in Myanmar and also in different countries. Moreover, this study would be useful for policy makers in drawing of sanitation and nutrition development program for malnutrition reduction among under five years old children.

### Limitations of study

Since the current study was a cross-sectional analytical study, further study with operational research or longitudinal cohort study design was recommended to provide the better understanding of the relationship between safely managed sanitation practices and childhood malnutrition.

## 5. Conclusion

There was high prevalence of stunting among children aged 5 to 59 months old children in Myanmar. Children who accessible to unsafely managed sanitation facilities was strongly associated with childhood stunting among under five children in Myanmar. And unproper waste water disposal practices and need times to collect drinking water were the WASH related influencing factors for stunting among under five children. Moreover, eating at least 4 diverse food items, maternal height, gender of children and vitamin A supplement were also significant factors for determining childhood stunting.

The following recommendations were made to enhance the nutritional status of children under the age of five. i) to promote the accessibility of the community’s safely managed sanitation facilities, ii) to provide technical training and knowledge sharing sessions to the community about household waste water management, iii) to promote the availability of drinking water at home, iv) to provide health education to mothers for the minimum diet diversity needs of under five children, v) to regularly provide Vitamin A supplement to 6-59 months old children, and vi) to conduct the further study with operational research or longitudinal cohort study design for understanding more the relationship between safely managed sanitation practices and childhood malnutrition.

### Ethic approval and consent to participate

This study protocol was approved by Khon Kaen University Ethics Committee for human research with the reference number HE652157. Mothers and fathers of the children provided the written informed consent.

## Data Availability

The full dataset presented in this study is available on request from the corresponding author.

## Acknowledgement

The author would like to express the sincere thanks to village and ward administers, community leaders and health staffs of study Townships for helping in data collection. And the author would like to thank all mothers and fathers of under-five children of study area for their participation and their commitment.

## Funds

No funding was received for this study from any funding agency in the public, commercial or not-for-profit sectors.

## Supplementary materials

Preformed questionnaires of “safely managed sanitation practice and childhood stunting among under five years old children in Myanmar” was attached.

